# U.S. Food and Drug Administration utilization of postmarketing requirements and postmarketing commitments, 2009-2018

**DOI:** 10.1101/2020.08.31.20184184

**Authors:** Joshua J. Skydel, Audrey D. Zhang, Sanket S. Dhruva, Joseph S. Ross, Joshua D. Wallach

## Abstract

**Background/Aims:** The U.S. Food and Drug Administration (FDA) outlines clinical studies as postmarketing requirements and commitments to be fulfilled following FDA approval of new drugs and biologics (“therapeutics”). As regulators have increasingly emphasized lifecycle evaluation of approved therapeutics, postmarketing studies are intended to advance our understanding of therapeutic safety and efficacy, yet little is known about how often clinical studies are outlined or the indications they investigate. Therefore, we characterized FDA postmarketing requirements and commitments for new therapeutics approved from 2009-2018.

**Methods:** We conducted a cross-sectional study of all novel therapeutics, including small molecule drugs and biologics, receiving original FDA approval from 2009-2018, using approval letters accessed through the Drug@FDA database. Outcomes included the number and characteristics of FDA postmarketing requirements and commitments for new therapeutics at original approval, including types of studies outlined, indications to be investigated, and clinical evidence to be generated.

**Results:** From 2009-2018, FDA approved 343 new therapeutics with 1978 postmarketing requirements and commitments. Overall, 750 (37.9%) postmarketing requirements and commitments outlined clinical studies. For 71 of 343 (20.7%) therapeutics, no postmarketing requirements nor commitments for clinical studies were outlined, while at least 1 was outlined for 272 (79.3%; median = 2 (IQR, 1-4)). Among these 272 therapeutics, the number of postmarketing requirements and commitments for clinical studies per therapeutic did not significantly change from 2009 (median 2 (IQR, 1-4)) to 2018 (median 2 (IQR, 1-3); P = .54). Among the 750 postmarketing requirements and commitments for clinical studies, 448 (59.7%) outlined new prospective cohort studies, registries, or clinical trials, while the remainder outlined retrospective studies, secondary analyses, or completion of ongoing studies. Although 455 (60.7%) clinical studies investigated only original approved therapeutic indications, 123 (16.4%) enrolled from an expansion of the approved disease population and 61 (8.1%) investigated diseases unrelated to approved indications.

**Conclusions:** Most therapeutics are approved by FDA with at least 1 postmarketing requirement or commitment for a clinical study, which outline investigations of safety or efficacy for both approved and unapproved indications. However, the median number of 2 clinical studies outlined has remained relatively constant over the last decade, despite increasing emphasis on lifecycle evaluation of approved therapeutics.

## Introduction

To receive regulatory approval by the U.S. Food and Drug Administration (FDA), new small molecule drugs and biologics (“therapeutics”) are generally required to be supported by two or more well-controlled studies demonstrating safety and efficacy.^1,2^ However, FDA has increasingly emphasized postmarket evidence generation in support of lifecycle evaluation of therapeutics.^3^ Furthermore, use of FDA’s expedited review programs,^4^ intended to speed the entry of therapeutics to market,^5,6^ has contributed to more therapeutics being approved on the basis of fewer pivotal trials,^7^ and trials using surrogate markers as primary endpoints.^8^ To generate evidence not available at therapeutic approval, FDA has the authority, under four statutes,^9^ to require sponsors to fulfill postmarketing requirements (PMRs) for clinical studies intended to generate safety data, confirm clinical benefit, or clarify the optimal use of therapeutics.^10^ FDA also collaborates with sponsors on voluntary postmarketing commitments (PMCs) to generate clinical evidence in support of ongoing therapeutic evaluation.^10^

While PMRs and PMCs represent an increasingly important source of safety and efficacy evidence for approved therapeutic indications, numerous analyses have noted shortcomings in their use.^11–13^ For instance, therapeutics are often approved without PMRs or PMCs.^14^ In such cases, sponsors rarely conduct clinical studies of original approved indications, instead investigating therapeutic uses for unapproved diseases (i.e. off-label) or expanded patient populations.^14^ These studies may support regulatory submissions for supplemental indication approvals, but can also promote off-label use of therapeutics without regulatory oversight of new clinical investigations.^15,16^ Previous analyses of new therapeutics approved from 2009 to 2012 found that PMRs and PMCs outlined at approval rarely require new prospective cohort studies, registries, or clinical trials, even though these are important sources of clinical evidence for understanding therapeutic efficacy and safety.^17,18^ Furthermore, new clinical studies from PMRs and PMCs are inconsistently completed and disseminated,^17–20^ despite generous timelines.^21^

Given the opportunity for PMRs and PMCs to promote the generation of clinical evidence for therapeutics, particularly with respect to safety and efficacy for original approved indications, it is important to understand the studies FDA has outlined in PMRs and PMCs, the indications they investigate, and whether they are intended to generate safety and/or efficacy data. Therefore, we characterized PMRs and PMCs outlined for therapeutics receiving original FDA approval from 2009 to 2018, an interval notable for FDA’s increasing emphasis on lifecycle evaluation and expanding use of expedited review programs,^3,7^ including the introduction of new programs.^4^ We assessed the utilization of each PMR or PMC authority, the types of studies outlined, and the indications for which PMRs and PMCs are anticipated to generate evidence.

## Methods

### Study Design and Sample

Three authors (J.J.S., A.D.Z., J.D.W.) used the publicly available Drugs@FDA database to identify all therapeutics that received original FDA approval from 1 January 2009 to 31 December 2018.^22^ We excluded generic drugs, reformulations and new combinations of previously approved therapeutics, and non-therapeutic agents (e.g., contrast agents), using previous methodology.^8^ Approval data were identified from FDA novel drugs summaries and related publications,^23–25^ including the type of application (New Drug Application vs. Biologic License Application) and whether the therapeutic underwent priority review, received accelerated approval pathway designation, or received orphan drug designation.

Using original FDA approval letters and drug labels, we abstracted the original FDA-approved indication(s) for each therapeutic. To define original approved indications, we recorded indicated disease(s) (e.g., inflammatory bowel disease, non-small cell lung cancer), disease characteristics (e.g., moderate-to-severe, metastatic), and treatment characteristics (e.g., second-line therapy, component of multi-drug regimen). Contraindications and other information relevant to therapeutic use was also collected. Indications were classified according to the World Health Organization Anatomic Therapeutic Classification system,^26^ collapsed into 7 categories (**Table 1**).

**Table 1.** Characteristics of 343 new therapeutics receiving original Food and Drug Administration approval, 2009-2018.

|  |  |  |  |  |
| --- | --- | --- | --- | --- |
| Therapeutic characteristic | Number of therapeutics | No. (%) |  | P values |
|  |  | No clinical PMRs or PMCs at approval<sup>a</sup> | At least one clinical PMR or PMC at approval |  |
| Total | 343 | 71 | 272 | - |
| Year of approval |  |  |  |  |
| 2009 | 26 (7.6) | 5 (7.0) | 21 (7.7) | .99 |
| 2010 | 21 (6.1) | 4 (5.6) | 17 (6.3) |  |
| 2011 | 28 (8.2) | 6 (8.5) | 22 (8.1) |  |
| 2012 | 36 (10.5) | 8 (11.3) | 28 (10.3) |  |
| 2013 | 24 (7.0) | 6 (8.5) | 18 (6.6) |  |
| 2014 | 39 (11.4) | 9 (12.7) | 30 (11.0) |  |
| 2015 | 45 (13.1) | 8 (11.3) | 37 (13.6) |  |
| 2016 | 20 (5.8) | 3 (4.2) | 17 (6.3) |  |
| 2017 | 45 (13.1) | 11 (15.5) | 34 (12.5) |  |
| 2018 | 59 (17.2) | 11 (15.5) | 48 (17.6) |  |
| Class |  |  |  |  |
| Drug | 258 (75.2) | 59 (83.1) | 199 (73.2) | .09 |
| Biologic | 85 (24.8) | 12 (16.9) | 73 (26.8) |  |
| Therapeutic area |  |  |  |  |
| Autoimmune, musculoskeletal, and dermatology | 39 (11.4) | 6 (8.5) | 33 (12.1) | < .001 |
| Cancer and hematology | 97 (28.3) | 16 (22.5) | 81 (29.8) |  |
| Cardiovascular and diabetes | 40 (11.7) | 10 (14.1) | 30 (11.0) |  |
| Gastrointestinal and metabolism | 31 (9.0) | 3 (4.2) | 28 (10.3) |  |
| Infectious disease | 55 (16.0) | 5 (7.0) | 50 (18.4) |  |
| Neurology and psychiatry | 34 (9.9) | 7 (9.9) | 27 (9.9) |  |
| Other | 47 (13.7) | 24 (33.8) | 23 (8.5) |  |
| Priority review |  |  |  |  |
| Yes | 185 (53.9) | 33 (46.5) | 152 (55.9) | .18 |
| No | 158 (46.1) | 38 (53.5) | 120 (44.1) |  |
| Accelerated approval |  |  |  |  |
| Yes | 41 (12.0) | 0 (0) | 41 (15.1) | < .001 |
| No | 302 (88.0) | 71 (100.0) | 231 (84.9) |  |
| Orphan drug designation |  |  |  |  |
| Yes | 142 (41.4) | 33 (46.5) | 109 (40.1) | .35 |
| No | 201 (58.6) | 38 (53.5) | 163 (59.9) |  |
| PMRs: postmarketing requirements; PMCs: postmarketing commitments. |  |  |  |  |
| <sup>a</sup> Includes therapeutics approved with no PMRs or PMCs and therapeutics approved with non-clinical PMRs and/or PMCs only. |  |  |  |  |

### Identifying postmarketing requirements and commitments

Building upon data collected for therapeutics receiving original FDA approval from 1 January 2009 to 31 December 2012,^17,18^ we used original FDA approval letters to identify all PMRs and PMCs outlined at the time of original approval for each therapeutic and the regulatory authority under which each was issued (**eTable 1 in Supplement**). PMRs and PMCs were categorized based on the type of study (e.g., new prospective, ongoing prospective, animal) described in approval letters, using previous methodology (**eBox in Supplement**).^17,18^ Any PMR or PMC describing a study with safety and/or efficacy outcomes (i.e., a clinical study) was considered a “clinical PMR or PMC”; all others were considered non-clinical. Clinical study categories included: new prospective cohort studies, registries, or clinical trials (“new prospective clinical studies”); completion of ongoing prospective clinical studies; new retrospective observational studies; or new analyses or follow-up for any clinical studies. Evidence generated by clinical PMRs and PMCs was characterized as safety, efficacy, or both.

### Characterizing indications of postmarketing requirements and commitments

We classified the indications for clinical PMRs and PMCs by comparing descriptions in approval letters of proposed postmarket studies to original FDA-approved therapeutic indications (**Table 2**). PMRs and PMCs were classified as generating evidence for the original approved disease (“original indication”), expanded disease populations beyond the scope of the original indication (“modified indication,” e.g., use in treatment-naïve patients when originally approved as second-line therapy for that disease), or new diseases not included in an original indication (“new indication”, e.g., chronic obstructive pulmonary disease when originally approved for the treatment of asthma). PMRs and PMCs outlining studies for original approved diseases were also classified as enrolling from the entirety of the indicated population (“general population”) or from a demographic (e.g., pediatric) or clinical (e.g., patients with comorbid chronic kidney disease) subgroup. Pediatric studies for original approved diseases were considered to investigate a demographic subgroup of the indicated population (**eAppendix in Supplement**).

**Table 2.** Sample classifications of indications for studies outlined in clinical postmarketing requirements and commitments.

|  |  |  |  |  |
| --- | --- | --- | --- | --- |
| **Therapeutic** | **Original FDA approved indication** | **PMR/PMC: authority;<sup>a</sup> description<sup>22</sup>** | **PMR/PMC study population** | **PMR/PMC indication classification<sup>b</sup>** |
| **Lurasidone** | Schizophrenia | PMC: 506B; “To evaluate the longer-term, i.e. maintenance, efficacy of lurasidone in the treatment of adults with schizophrenia...” | Adult patients with schizophrenia | Original indication, general population |
| **Simeprevir** | Chronic hepatitis C infection, as a component of a combination antiviral treatment regimen | PMC: 506B; “Submit the final report and datasets for trial... [in] Subjects who are Co-Infected with Human Immunodeficiency Virus Type 1 (HIV-1).” | Chronic hepatitis C infection, with HIV-1 coinfection | Original indication, clinical subgroup |
| **Linagliptin** | Type 2 diabetes mellitus | PMR: PREA; “...evaluate efficacy, safety, and pharmacokinetics of linagliptin for the treatment of type 2 diabetes mellitus in pediatric patients ages 10 to 16 years...” | Pediatric patients with type 2 diabetes mellitus | Original indication, demographic subgroup |
| **Cabazitaxel** | Hormone-refractory metastatic prostate cancer, in combination with prednisone, for patients previously treated with a docetaxel-containing treatment regimen | PMR: FDAAA; “Conduct a Phase 3 randomized controlled trial in patients with hormone-refractory metastatic prostate cancer...with prednisone as first-line therapy.” | First-line therapy with prednisone for (i.e., previously untreated) patients with hormone-refractory metastatic prostate cancer | Modified indication |
| **IncobotulinumtoxinA** | Cervical dystonia and blepharospasm | PMC: 506B; “Randomized, double-blind, adequate and well controlled, multiple fixed-dose, parallel group clinical trial... [in] adults with lower extremity spasticity.” | Adult patients with lower extremity spasticity | New indication |
| PMR: postmarketing requirement; PMC: postmarketing commitment; FDAAA: Food and Drug Administration Amendments Act; PREA: Pediatric Research Equity Act.<br><sup>a</sup> “506B” refers to PMCs subject to reporting requirements under section 506B of the Federal Food, Drug, and Cosmetic Act.<br><sup>b</sup> Additional example PMR/PMC classifications are included in eAppendix 1. |  |  |  |  |

Clinical PMRs and PMCs were abstracted using a prespecified algorithm, using descriptions from FDA approval letters supplemented as necessary by corresponding study registrations on ClinicalTrials.gov to abstract study design, indication, and safety and/or efficacy endpoints. PMRs and PMCs generating evidence for both original and modified or new indications were classified as “modified” or “new indication,” as applicable. PMRs and PMCs were abstracted by one author (J.J.S.), with uncertainties resolved via consensus amongst all authors. Another author (J.D.W.) validated abstractions using a 20% random sample of PMRs and PMCs. Analyses were conducted from 29 July 2019 to 23 March 2020.

### Statistical Analysis

We used descriptive statistics to summarize new therapeutics receiving FDA approval from 2009 to 2018 and to characterize PMRs and PMCs by FDA, including characteristics such as issuing authority, study type, investigated indications, and evidence generated. We used Fisher’s exact and Kruskal-Wallis tests to evaluate associations between therapeutic characteristics and the issuance of PMRs and PMCs. All statistical tests were 2-sided, significance was set at 0.05, and analyses were performed using R (version 3.5.1).

### Ethical Review and Reporting Guideline

This study was conducted using publicly available, nonclinical data and did not require institutional review board approval. It adheres to the Strengthening the Reporting of Observational Studies in Epidemiology (STROBE) reporting guideline for cross-sectional studies.

## Results

### Characteristics of new therapeutics

From 2009 to 2018, FDA approved a total of 356 new therapeutics for 388 original indications. After excluding ineligible therapeutics, there were 343 (96.3%) therapeutics approved for 375 original indications included in our analyses (**Table 1**). Among the 343 therapeutics, 258 (75.2%) were small molecule drugs and 85 (24.8%) were biologics; 185 (53.9%) underwent priority review, 41 (12.0%) received accelerated approval designation, and 142 (41.4%) were granted orphan designation. The most frequently represented therapeutic area was cancer or hematologic disease (97, 28.3%).

A total of 311 (90.7%) therapeutics were approved with at least 1 PMR or PMC. For 272 (79.3%) therapeutics, at least 1 clinical PMR or PMC was outlined, while none were outlined for 71 (20.7%) therapeutics. All 41 therapeutics granted accelerated approval designation were approved with at least 1 clinical PMR or PMC. Therapeutics approved with at least 1 clinical PMR or PMC were more likely to have been granted accelerated approval designation when compared with therapeutics approved without clinical PMRs or PMCs (P < .001), and there were differences by therapeutic area (P < .001).

### Postmarketing requirements and commitments for new therapeutics

We identified a total of 1978 PMRs and PMCs for therapeutics receiving original FDA approval from 2009 to 2018, including 1123 (56.8%) PMRs and 855 (43.2%) PMCs. There were 1228 (62.1%) PMRs and PMCs outlining non-clinical studies and 750 (37.9%) outlining clinical studies (**eTable 2 in Supplement**). Among the 750 clinical studies, four-fifths (600/750, 80.0%) were outlined in PMRs and one-fifth (150/750, 20.0%) in PMCs. Most clinical PMRs and PMCs (448/750, 59.7%) outlined new prospective clinical studies (i.e., prospective cohort studies (48/750, 6.4%), registries (45/750, 6.0%), or clinical trials (355/750, 47.3%)); 125 of 750 (16.7%) outlined the completion or submission of results from ongoing prospective clinical studies. Three-quarters of PMRs issued under PREA (191/257, 74.3%) and one-half issued under accelerated approval (31/62, 50.0%) outlined new prospective clinical studies. Despite representing over one-quarter of new therapeutics, cancer and hematology products only had 71 (71/448, 15.8%) PMRs and PMCs for new prospective clinical studies (**eTable 3 in Supplement**).

The median number of PMRs and PMCs for new therapeutics overall was 5 (interquartile range (IQR), 2-8), and the median number of clinical PMRs and PMCs was 2 (IQR, 1-3). There was a non-significant decrease in the median number of clinical PMRs and PMCs for new therapeutics overall from 2009 (2, IQR, 1-2) to 2018 (1, IQR, 1-3) (**Figure 1**, P = .54). Among therapeutics approved with at least 1 clinical PMR or PMC, the median number of clinical PMRs and PMCs was 2 for most years from 2009 (2, IQR,1-4) to 2018 (2, IQR, 1-3).

**Figure 1.**
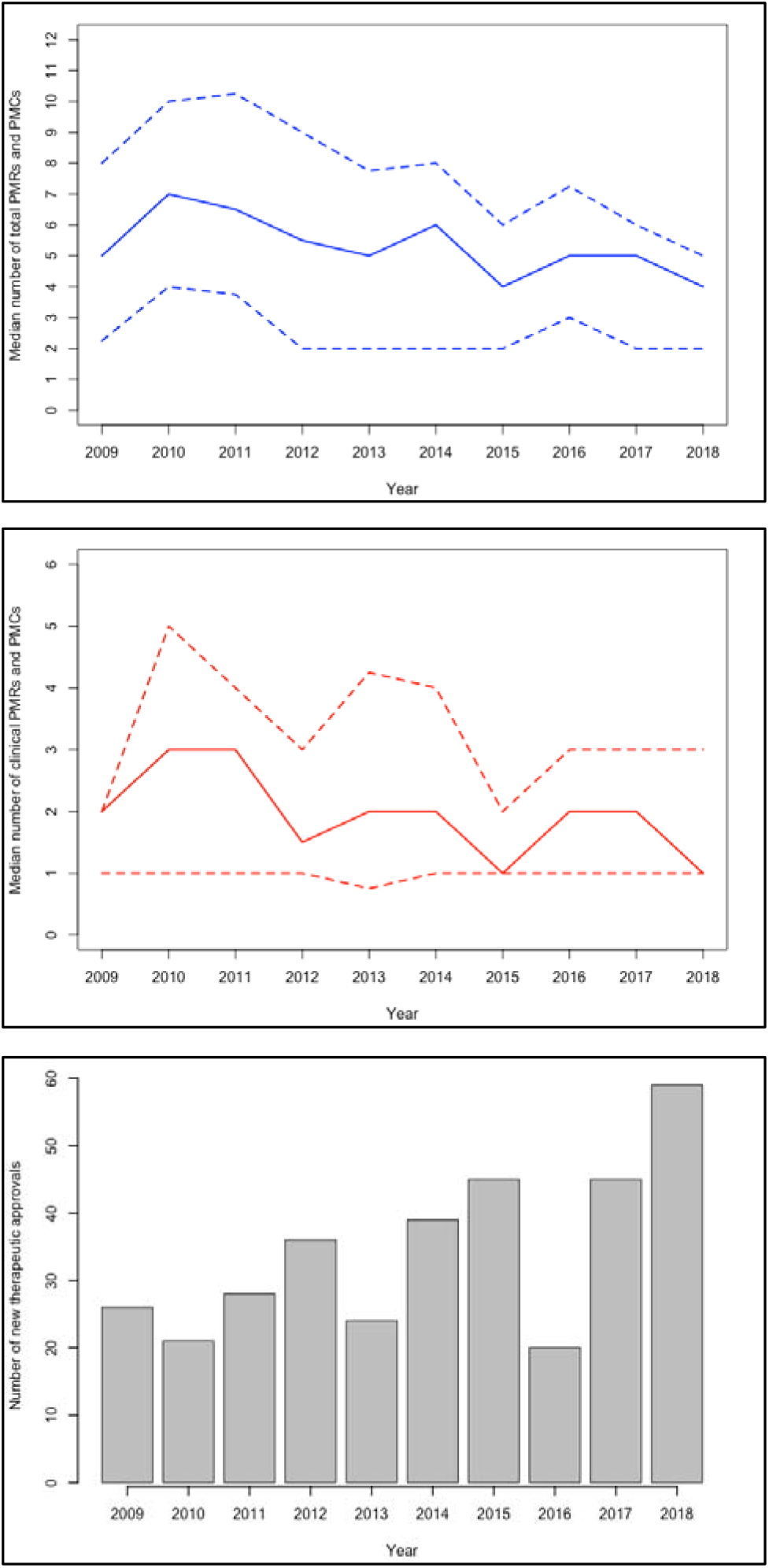
Postmarketing requirements and commitments outlined for therapeutics receiving original Food and Drug Administration approval, 2009-2018. (a) Median (solid line; interquartile range between dashed lines) total number of postmarketing requirements and commitments outlined for new therapeutics, 2009-2018. (b) Median (solid line; interquartile range between dashed lines) number of clinical postmarketing requirements and commitments outlined for new therapeutics, 2009-2018. (c) Number of new therapeutic approvals by US Food and Drug Administration, 2009-2018.

### Characteristics of clinical postmarketing requirements and commitments

Most clinical PMRs and PMCs outlined safety endpoints to be evaluated, either with (314/750, 41.9%) or without (330/750, 44.0%) additional efficacy endpoints (**Table 3**). Efficacy endpoints were outlined in 420 of 750 (56.0%) clinical PMRs and PMCs. Among 330 clinical PMRs and PMCs specifying only safety endpoints, over three-fourths were issued under FDAAA (258/330, 78.2%). However, more than one-fifth of the 331 PMRs issued under FDAAA included an efficacy endpoint (73/331, 22.1%).

**Table 3.** Characteristics of clinical postmarketing requirements and commitments for new therapeutics receiving original Food and Drug Administration approval, 2009-2018.

| PMR or PMC authority, No. (%) |  | Total clinical PMRs or PMCs<sup>b</sup> | PMR/PMC study indication |  |  |  |  | Study design |  | Study objective |  |  |
| --- | --- | --- | --- | --- | --- | --- | --- | --- | --- | --- | --- | --- |
|  |  |  | Original indication |  | Modified indication | New indication | Unclear<sup>d</sup> | Trial | Observational | Safety | Efficacy | Safety and Efficacy |
|  |  |  | General population | Subgroup<sup>c</sup> |  |  |  |  |  |  |  |  |
| **Total** |  | 750 | 150 (20.0) | 305 (40.7) | 123 (16.4) | 61 (8.1) | 111 (14.8) | 576 (76.8) | 174 (23.2) | 330 (44.0) | 106 (14.1) | 314 (41.9) |
| **PMR** | **FDAAA** | 331 | 94 (28.4) | 61 (18.4) | 48 (14.5) | 23 (6.9) | 105 (31.7) | 173 (52.3) | 158 (47.7) | 258 (77.9) | 8 (2.4) | 65 (19.6) |
|  | **PREA** | 207 | 0 (0) | 166 (80.2) | 19 (9.2) | 18 (8.7) | 4 (1.9) | 207 (100) | 0 (0) | 64 (30.9) | 3 (1.4) | 140 (67.6) |
|  | **AA** | 59 | 19 (32.2) | 10 (16.9) | 24 (40.7) | 6 (10.2) | 0 (0) | 58 (98.3) | 1 (1.7) | 1 (1.7) | 34 (57.6) | 24 (40.7) |
|  | **AER** | 3 | 3 (100.0) | 0 (0) | 0 (0) | 0 (0) | 0 (0) | 0 (0) | 3 (100.0) | 0 (0) | 0 (0) | 3 (100.0) |
| **PMC** | **506B<sup>a</sup>** | 141 | 34 (24.1) | 66 (46.8) | 29 (20.6) | 10 (7.1) | 2 (1.4) | 129 (91.5) | 12 (8.5) | 7 (5.0) | 58 (41.1) | 76 (53.9) |
|  | **Non-506B<sup>a</sup>** | 9 | 0 (0) | 2 (22.2) | 3 (33.3) | 4 (44.4) | 0 (0) | 9 (100.0) | 0 (0) | 0 (0) | 3 (33.3) | 6 (66.7) |

PMR: postmarketing requirement; PMC: postmarketing commitment; FDAAA: Food and Drug Administration Amendments Act; PREA: Pediatric Research Equity Act; AA: Accelerated Approval; AER: Animal Efficacy Rule.
<sup>a</sup>“506B” refers to PMCs subject to reporting requirements under section 506B of the Federal Food, Drug, and Cosmetic Act. PMCs not subject to this rule are denoted as “Non-506B” PMCs.
<sup>b</sup>“Clinical PMRs/PMCs” include those outlining new prospective cohort studies, registries, or clinical trials; completion or submission of results from ongoing prospective clinical studies; new retrospective observational studies; or new analysis, follow-up, or flexible analyses of clinical studies.
<sup>c</sup> PMRs and PMCs generating evidence on subgroups of the original indication may evaluate demographic subgroups, clinical subgroups, or both.
<sup>d</sup>“Unclear” indications were those for which a defined disease population could not be determined from PMR or PMC description and, if identified, corresponding study registration on ClinicalTrials.gov.

The majority of the 750 clinical PMRs and PMCs were intended to generate evidence for original approved indications, either for the general disease population (150/750, 20.0%) or for a clinical and/or demographic subgroup (305/750, 40.7%) (**Table 2**). However, nearly one-quarter (184/750, 24.5%) of clinical PMRs and PMCs described studies of unapproved indications, including 123 (123/184, 66.8%) evaluating modified indications expanding therapeutic uses within original disease populations (**Table 4**). A total of 61 (61/184, 33.2%) clinical PMRs and PMCs evaluated therapeutic uses for unapproved diseases unrelated to original indications, including approximately one-third (21/61, 34.4%) generating preliminary safety and/or efficacy data in patients with nonspecific diagnoses such as “solid tumors” or “bacterial infections.” Clinical PMRs and PMCs for modified or new indications most often outlined new prospective clinical studies (103/184, 56.0%), of which nearly one-quarter (24/103, 23.3%) investigated therapeutic uses unrelated to original indications. For clinical PMRs issued under the accelerated approval authority, 30 of 59 (50.8%) outlined clinical studies generating evidence on modified or new therapeutic indications (**Table 3**).

**Table 4.** Clinical postmarketing requirements and commitments investigating modified or new therapeutic indications.

|  |  |  |  |  |  |
| --- | --- | --- | --- | --- | --- |
| **Study type, No. (%)** | **Clinical PMRs and PMCs investigating modified or new indications** | **Clinical PMRs and PMCs investigating modified indications** | **Clinical PMRs and PMCs investigating new indications<sup>a</sup>** |  |  |
|  |  |  | **Total** | **New therapeutic use<sup>b</sup>** | **Clinical safety and efficacy data<sup>b</sup>** |
| **Total** | 184 | 123 (66.8) | 61 (33.2) | 40 (65.6) | 21 (34.4) |
| **New prospective cohort study, registry, or clinical trial** | 103 | 65 (63.1) | 38 (36.9) | 24 (63.2) | 14 (36.8) |
| **Complete or submit results from prospective cohort study, registry, or clinical trial** | 41 | 30 (73.2) | 11 (26.8) | 6 (54.5) | 5 (45.5) |
| **New retrospective observational study** | 2 | 1 (50.0) | 1 (50.0) | 1 (100.0) | 0 (0) |
| **New analysis or follow-up for cohort study, registry, or clinical trial, or “flexible” requirements** | 38 | 27 (71.1) | 11 (28.9) | 9 (81.8) | 2 (18.2) |
| PMR: postmarketing requirement; PMC: postmarketing commitment.<br><sup>a</sup> Clinical PMRs and PMCs were considered to investigate a new indication when the outlined study was to be conducted in a disease population not related to the original approved indication. This included studies of unapproved therapeutic uses in new disease populations as well as studies intended to generate basic clinical data in a nonspecific disease population (e.g., patients with “solid tumors” or “bacterial infections”).<br><sup>b</sup> Values in parentheses reflect percentages of clinical PMRs and PMCs investigating new therapeutic indications. |  |  |  |  |  |

## Discussion

Among 1978 PMRs and PMCs outlined for 343 therapeutics originally approved by FDA from 2009 to 2018, we found variation in the number, design, and characteristics of clinical studies they described. Just under 40% of all PMRs and PMCs outlined clinical studies, and even fewer described new prospective cohort studies, registries, or clinical trials to be conducted in the postmarket period. Although the majority of new therapeutics were approved with at least 1 clinical PMR or PMC, the median number of clinical PMRs and/or PMCs outlined for those therapeutics was 2, and was relatively consistent from 2009 to 2018. Clinical studies outlined in PMRs and PMCs were frequently intended to generate safety and efficacy evidence for approved indications, but nearly one-quarter of clinical PMRs and PMCs described studies with the potential to generate evidence on therapeutic uses not encompassed by their original approved indications. These findings suggest that a greater number of PMRs and PMCs outlining clinical studies of approved indications may be needed to address evidentiary gaps and inform clinical decision making.

Despite FDA’s increasing emphasis on lifecycle evaluation,^3^ we did not identify a change in the number of clinical PMRs and PMCs outlined for new therapeutics over the last decade, with fewer than one-quarter of PMRs and PMCs outlining new prospective studies such as clinical trials. Between 2009 and 2018, the median number of clinical PMRs and PMCs outlined at therapeutic approval remained 2 for therapeutics approved with at least 1 clinical PMR or PMC and decreased non-significantly from 2 to 1 for therapeutics overall. This occurred in the context of therapeutic approvals increasingly being based on fewer pivotal trials,^7^ often using surrogate endpoints.^27^ Use of FDA’s expedited review programs is increasing,^4^ including the breakthrough therapy designation implemented in 2012, which supports the approval of new therapeutics considered promising on the basis of preliminary clinical evidence.^6^ These programs reduce the amount of clinical evidence available for new therapeutics at approval,^28^ yet the minority of PMRs and PMCs outlined in the previous decade are for new prospective clinical studies. Together, these findings suggest that clinical PMRs and PMCs may not fully compensate for decreasing numbers of premarket clinical trials for new therapeutics. Expanded use of PMRs and PMCs to outline prospective clinical studies may represent the most effective approach to supplement decreasing numbers of premarket clinical trials and generate postmarket evidence to inform clinical decision making.

We found that clinical PMRs and PMCs frequently were expected to address both safety and efficacy endpoints, possibly reflecting FDA’s vision for PMRs and PMCs as flexible responses to clinical questions arising at the time of or following approval.^29^ However, nearly one-quarter of clinical PMRs and PMCs focused on modified or unapproved therapeutic indications, including one-half of confirmatory PMRs for therapeutics receiving accelerated approval designation. Previous studies have noted that postmarket clinical studies by industry sponsors frequently evaluate new therapeutic uses,^14^ while others have suggested they play a role in promoting medication use after approval.^30^ This has been observed for therapeutics receiving accelerated approval designation, which are integrated into clinical practice, including new applications, without confirmation of clinical benefit for original indications.^31^ We also identified PMRs and PMCs describing new prospective clinical studies potentially investigating modified or new indications. For example, ofatumumab originally received FDA accelerated approval for the treatment of chronic lymphocytic leukemia (CLL) refractory to alemtuzumab and fludarabine. However, a confirmatory PMR outlined under the accelerated approval authority required completion of a clinical trial of ofatumumab in previously untreated patients with CLL, potentially expanding its indication to include patients for which there was still a need for confirmatory evidence of efficacy at original approval. Similarly, for deferiprone, indicated for the treatment of transfusional iron overload in patients with thalassemia syndromes, a PMR outlined under the accelerated approval authority investigated use in patients with sickle cell disease, for which current pharmacologic management is limited.^32^ While PMRs and PMCs investigating novel indications may inform the use of therapeutics in patients with more severe or earlier stage disease, or even distinct diseases, they will not generate efficacy evidence to inform decisions to treat the originally approved indications.

Accelerating therapeutic approvals represent a tradeoff between the benefit to patients of faster access to novel therapies and the need for comprehensive evidence demonstrating safety and efficacy.^33^ Following therapeutic approval, adverse event reports, electronic health records, and insurance claims allow FDA to monitor therapeutic use and identify safety signals requiring communication to the public or regulatory action.^34,35^ However, there are shortcomings to real world data sources.^36,37^ A report by the U.S. Office of the Inspector General noted that PMRs frequently result in labeling changes and other actions by FDA to support therapeutic safety, suggesting their importance for generating evidence of value to clinical practice.^38^ PMRs and PMCs enable FDA to target evidentiary shortcomings, and opportunities exist to use them in coordination with real world evidence to refine assessments of therapeutic safety and efficacy and support FDA’s transition to lifecycle evaluation, such as through the conduct of pragmatic clinical trials. FDA has an opportunity to outline a greater number of new prospective clinical studies in PMRs and PMCs for new therapeutics, advancing our understanding of their optimal uses as they are integrated into clinical practice. While PMRs and PMCs can be used to investigate expanded or new therapeutic uses, these should not replace studies generating clinical evidence for original indications.

## Limitations

Our study has several limitations. First, we abstracted PMRs and PMCs based on descriptions in original approval letters, which are sometimes too brief to comprehensively characterize study indications or design elements, such as enrollment or trial duration.^17^ Although we used study registrations on ClinicalTrials.gov to supplement PMR and PMC descriptions, these were nearly always, but not universally, available. Second, we characterized indications based on publicly available data, and therefore may not have captured FDA’s objectives for some PMRs and PMCs. However, in the absence of additional information from FDA, PMR and PMC descriptions represent the best public resource for characterizing these postmarket studies. Third, we did not evaluate the completion of or reporting of results from clinical PMRs and PMCs, which previous studies have suggested takes place for only approximately 50% of clinical PMR and PMCs.^17–19^ Further analyses may provide additional information about how often postmarket clinical evidence becomes available to clinicians and patients. Lastly, we limited our analyses to PMRs and PMCs from 10 years of new therapeutic approvals. Although postmarketing studies were outlined prior to 2008, the term “postmarketing study commitments” referred to both required and agreed-upon studies, making it difficult to differentiate PMRs from PMCs.^10^ Our sample represents, to our knowledge, the largest analysis of PMRs and PMCs since terminology was standardized.

## Conclusions

Among 343 therapeutics that received original FDA approval from 2009 to 2018, most were approved with at least 1 clinical PMR or PMC. However, the median number of studies investigating therapeutic safety and/or efficacy was 2 per approval, fewer than one-quarter outlined new prospective clinical studies, and studies investigated both approved and unapproved indications. Given FDA’s commitment to expedited approval and increasing emphasis of lifecycle therapeutic evaluation, a greater number of PMRs and PMCs outlining clinical studies of approved indications may be needed to address evidentiary gaps and inform clinical decision making.

## Data Availability

Data will be shared on osf.io upon publication.

## Acknowledgments

*Author contributions:* Mr. Skydel and Dr. Wallach had full access to all the data in the study and take responsibility for the integrity of the data and the accuracy of the data analysis.

*Concept and design:* Skydel, Dhruva, Ross, Wallach.

*Acquisition, analysis, or interpretation:* All authors.

*Drafting of the manuscript:* Skydel.

*Critical revision of the manuscript for important intellectual content:* All authors.

*Statistical analysis:* Skydel.

*Supervision:* Wallach.

*Funding/support and role of the sponsor:* This project was not supported by any external grants or funds.

*Conflicts of interest disclosure:* In the past 36 months, JDW received research support through the Collaboration for Research Integrity and Transparency from the Laura and John Arnold Foundation and through the Center for Excellence in Regulatory Science and Innovation (CERSI) at Yale University and the Mayo Clinic (U01FD005938). JSR is a former Associate Editor of JAMA Internal Medicine, a current Research Editor at BMJ and received research support through Yale from Johnson and Johnson to develop methods of clinical trial data sharing, from Medtronic, Inc. and the Food and Drug Administration (FDA) to develop methods for postmarket surveillance of medical devices (U01FD004585), from the Centers of Medicare and Medicaid Services (CMS) to develop and maintain performance measures that are used for public reporting, from the FDA to establish a Center for Excellence in Regulatory Science and Innovation (CERSI) at Yale University and the Mayo Clinic (U01FD005938), from the Blue Cross Blue Shield Association to better understand medical technology evaluation, from the Agency for Healthcare Research and Quality (R01HS022882), and from the Laura and John Arnold Foundation. SSD received support as a Scholar in the Yale University / Mayo Clinic FDA CERSI, from the Greenwall Foundation, and from the National Institutes of Health / National Heart, Lung, and Blood Institute K12HL138046.

*Data sharing:* Data will be shared on osf.io upon publication.

